# Birth during the COVID-19 pandemic, but not maternal SARS-CoV-2 infection during pregnancy, is associated with lower neurodevelopmental scores at 6-months

**DOI:** 10.1101/2021.07.12.21260365

**Authors:** Lauren C. Shuffrey, Morgan R. Firestein, Margaret Kyle, Andrea Fields, Carmela Alcántara, Dima Amso, Judy Austin, Jennifer M. Bain, Jennifer Barbosa, Mary Bence, Catherine Bianco, Cristina Fernández, Sylvie Goldman, Cynthia Gyamfi-Bannerman, Violet Hott, Yunzhe Hu, Maha Hussain, Pam Factor-Litvak, Maristella Lucchini, Arthur Mandel, Rachel Marsh, Danielle McBrian, Mirella Mourad, Rebecca Muhle, Kimberly Noble, Anna Penn, Cynthia Rodriguez, Ayesha Sania, Wendy G. Silver, Kally C. O’Reilly, Melissa Stockwell, Nim Tottenham, Martha G. Welch, Noelia Zork, William P. Fifer, Catherine Monk, Dani Dumitriu

## Abstract

The intrauterine environment strongly influences development. Neurodevelopmental effects of *in utero* exposure to maternal SARS-CoV-2 infection are widely speculated but currently unknown. The COVID-19 Mother Baby Outcomes (COMBO) initiative was established at Columbia University Irving Medical Center (CUIMC) in New York City to prospectively study the health and wellbeing of infants with and without *in utero* exposure to maternal SARS-CoV-2 infection. We report findings on 6-month neurodevelopmental outcomes using the parental-report *Ages & Stages Questionnaire, 3^rd^ Edition* (ASQ-3), from 107 *in utero* exposed and 131 unexposed full-term infants born between March and December, 2020. We compare these infants to a historical cohort comprised of 62 infants born at CUIMC at least two months prior to the onset of the pandemic. *In utero* exposure to maternal SARS-CoV-2 infection was not associated with differences on any ASQ-3 subdomain regardless of infection timing or severity, however, infants born during the pandemic had significantly lower scores on gross motor, fine motor, and personal-social subdomains when compared to the historical cohort. Infants born to women who were in the first trimester of pregnancy during the peak of the pandemic in NYC had the lowest personal-social scores. Birth during the pandemic, but not maternal SARS-CoV-2 infection, was associated with differences in neurodevelopmental outcomes at 6-months. These early findings suggest significantly higher public health impact for the generation born during the COVID-19 pandemic than previously anticipated.

## Introduction

Globally, over 200 million infants have been born since the onset of the COVID-19 pandemic. During the first two weeks of universal testing at the height of the pandemic in New York City (NYC), 14% of laboring women in the Columbia University Irving Medical Center (CUIMC) hospital system tested positive for severe acute respiratory syndrome coronavirus 2 (SARS-CoV-2) by nasopharyngeal polymerase chain reaction (PCR)^1^. To date, 2% of the world’s population has been infected at some point during the pandemic. While impossible to precisely quantify, even the most conservative estimates of the total number of infants worldwide with *in utero* exposures to maternal SARS-CoV-2 infection range in the millions.

Fetal exposure to perturbations of the intrauterine environment is implicated in altered brain development and long-term offspring vulnerability for neurodevelopmental and psychiatric sequelae^2-15^. Although vertical transmission of SARS-CoV-2 from mother to fetus is exceedingly rare^16-20^, data from prior human coronavirus outbreaks (severe acute respiratory syndrome (SARS), Middle East respiratory syndrome (MERS)) suggest that severe infection during pregnancy may not only impact maternal health^21^, but may also increase risk for several adverse infant outcomes through mechanisms related to maternal immune activation^22^. Other viral illnesses during pregnancy are associated with higher risk for neurodevelopmental deficits, including motor delays^23^, as in the case of *in utero* HIV-exposed uninfected infants^5-15^. Additional epidemiological support for this association comes from naturalistic studies of viral epidemics that identified population-level impacts. Cohort studies of the generation born during the 1918 H1N1 pandemic found lower child educational attainment and adult socioeconomic status^24^. The 1964 rubella pandemic led to a 10-15-fold increase in autism spectrum disorder or schizophrenia in offspring^25,26^. There is a critical need to determine the neurodevelopmental effects of fetal exposure to maternal SARS-CoV-2 infection^27-31^, especially given the well-established benefits of early identification of at-risk children^32-34^.

The COVID-19 Mother Baby Outcomes (COMBO) Initiative is a prospective matched case-control study established at CUIMC in Spring 2020 to examine the effects of *in utero* exposure to maternal SARS-CoV-2 infection on the health and wellbeing of both mother and child living in the first U.S. pandemic epicenter (www.ps.columbia.edu/COMBO). Based on evidence from prior studies^23,35,36^, we hypothesized maternal SARS-CoV-2 infection during pregnancy would be associated with delays in social and motor development at 6-months of age. Additionally, we compared infants born during the COVID-19 pandemic to a historical cohort born at the same medical center using the same neurodevelopmental assessment.

## Participants and Methods

### Study design and participants

This analysis includes infants enrolled in the COMBO Initiative born between March and December, 2020, and an available historical comparison cohort of infants born at the same institution between November, 2017 and January, 2020. Data using the same 6-month postnatal neurodevelopmental measures were available for participants enrolled in both of these cohorts. Preterm delivery (gestational age (GA) <37 weeks) was an exclusionary criteria for the historical cohort; therefore, only full-term COMBO infants were included.

All mother-infant dyads participating in the COMBO Initiative received prenatal care and delivered at the CUIMC-affiliated NewYork-Presbyterian (NYP) Morgan Stanley Children’s Hospital (MSCH) or NYP Allen Pavilion Hospital since the onset of the pandemic (first NYC laboring woman diagnosed 3/13/2020 at MSCH). All dyads with a documented SARS-CoV-2 infection during pregnancy (exposed) according to the electronic health record (EHR) were invited to participate and were enrolled during pregnancy or in the first few months postpartum. For each exposed dyad, 1-3 unexposed dyads, defined as the absence of EHR documentation of maternal SARS-CoV-2 infection during pregnancy and at delivery, were matched based on infant sex, GA at birth, mode of delivery, and date of birth (DOB) within a two-week window. Infants included in the historical cohort were recruited from the Well Baby Nursery at MSCH as part of a separate protocol. Study procedures were approved by the CUIMC Institutional Review Board (IRB) for the COMBO cohort and by the New York State Psychiatric Institute IRB for the historical cohort and informed consent was obtained from all participants.

Data at 6-months of age were available for 248 full-term infants enrolled in the pandemic cohort and 71 infants enrolled in the historical cohort. Data were excluded from 10 pandemic cohort and 9 historical cohort infants due to completion of the neurodevelopmental assessment outside of the eligibility window. The final sample included data from 300 infants: n=238 in the pandemic cohort (n=107 exposed; n=131 unexposed) and n=62 in the historical cohort.

### Determination of SARS-CoV-2 infection status

NYP’s hospital practices during the COVID-19 pandemic have been published^37^. Mother-infant separation, which is known to affect neurodevelopmental outcomes^38^, was never implemented. The labor and delivery (L&D) units of the NYP hospital system implemented universal SARS-CoV-2 testing of all delivering patients by nasopharyngeal PCR on 3/22/2020 and by serological testing for antibodies on 7/20/2020. Additional symptom-based testing occurred throughout pregnancy and results obtained from external testing sites were recorded in the EHR when possible. For each mother with a positive SARS-CoV-2 PCR and/or serology test during pregnancy (defined based on last menstrual period and delivery date) identified by automated EHR extraction, a careful chart review was conducted to determine the date of symptom onset, which was then referenced to infant GA to determine the trimester of *in utero* exposure. Exact exposure timing was determined for 79 of 107 mothers (74%) included in this analysis. For 28 (26%) asymptomatic serology-positive mothers, an exact date of symptom onset could not be determined, so the date with the peak number of SARS-CoV-2 infections during the first wave of the pandemic in NYC (4/6/2020) was used to impute the trimester of exposure. SARS-CoV-2 infection severity was categorized as asymptomatic, mild (defined as nonpneumonia or mild pneumonia) or severe (defined as dyspnea, respiratory rate ≥30/min, blood oxygen saturation≤93%, partial pressure of arterial oxygen to fraction of inspired oxygen ratio<300, and/or lung infiltrates>50% within 24-48 hours^16^).

Based on EHR and/or maternal-report, five infants in the pandemic cohort and zero infants in the historical cohort were infected with SARS-CoV-2 between birth and the neurodevelopmental assessment.

### Assessment of infant 6-month neurodevelopment

Between 5 months, 0 days, and 6 months, 30 days (186.79±11.75 days), infant neurodevelopment was assessed by maternal-report using the *Ages & Stages Questionnaire, 3^rd^ Edition* (ASQ-3) in English or Spanish via secure REDCap survey. The ASQ-3 was completed between October, 2020 and June, 2021 for the pandemic cohort and between May, 2018 and July, 2020 for the historical cohort. The ASQ-3 is a validated, widely used, standardized, level 1 screening tool based on parental-report that reliably assesses five key developmental domains: communication, fine and gross motor, problem solving, and personal-social skills. The ASQ-3 has strong concurrent validity (r=0.85), two-week test-retest reliability (r=0.75-0.82), inter-observer reliability (r=0.43-0.69), and internal consistency (α=0.51-0.87)^39,40^.

### Statistical analyses

Statistical analyses were conducted in R version 4.0.2. The experimental design (forced-choice online survey and chart review) resulted in no missingness. Power analyses were based on the primary outcome investigating mean differences in ASQ-3 subdomain scores between infants with and without *in utero* exposure to maternal SARS-CoV-2 infection using analyses of covariance (ANCOVAs), for which we had 90% power to detect a 0.5 SD difference in each subdomain using a two-sided test with alpha±0.05 (see Supplementary Materials). Within the exposed subset, we also examined the effect of SARS-CoV-2 timing and severity on ASQ-3 subdomains via ANCOVA. For secondary analyses, ANCOVAs were used to determine mean differences in ASQ-3 subdomains between the pandemic and historical cohorts. For both primary and secondary analyses, we implemented minimally (GA at birth, infant sex, and infant age at assessment) and fully adjusted models (additionally controlled for maternal race, ethnicity, age at delivery, educational attainment, parity, and mode of delivery). Percentages of infants screening below cutoffs indicative of delays per ASQ-3 developers^39^ were computed for each subdomain and for overall ASQ-3. Fisher’s Exact Test was used to examine group differences in screening positive for delays between the pandemic and historical cohorts. For ASQ-3 subdomains where a significant main effect was identified, tertiary analyses examined scores in relation to trimester of pregnancy during peak NYC SARS-CoV-2 cases (4/6/2020) using fully adjusted ANCOVA models with general linear hypothesis testing using Tukey pairwise contrasts.

Mean differences with confidence intervals (95% CI) are reported for all significant main effects. Estimated marginal means are reported in supplemental tables. Significance was set at p<0.05.

## Results

### Cohort characteristics

Highlighting the disproportionate impact of COVID-19 on already medically disadvantaged groups, SARS-CoV-2 infected mothers significantly differed from non-infected mothers in their self-reported race (X^2^=18.33, p=0.005), ethnicity (X^2^=21.07, p<0.001), and education attainment (X^2^=21.80, p=0.001), with a higher proportion identifying as other or mixed race (34.6% versus 21.4%) and Hispanic or Latino/a/x or Spanish ethnicity (64.5% versus 38.9%) and a lower proportion reporting a graduate degree (16.8% versus 40.5%) (Table 1). Compared to the pandemic cohort, the historical cohort significantly differed in race (X^2^=51.04, p<0.001), ethnicity (X^2^=37.85, p<0.001), education attainment (X^2^=35.17, p<0.001), and mode of delivery (X^2^=16.95, p<0.001). Group differences were accounted for in all fully adjusted models.

**Table 1.** Cohort characteristics.

| Table 1. Cohort characteristics |  |  |  |  |  |  |  |  |  |
| --- | --- | --- | --- | --- | --- | --- | --- | --- | --- |
| Variable<br>No. (%) or Median (Min, Max) | Overall<br>Sample<br>(N=300) | SARS-CoV-2<br>Unexposed<br>(N=131) | SARS-CoV-2<br>Exposed<br>(N=107) | SARS-CoV-2 Unexposed<br>vs. Exposed |  | Historical<br>Cohort<br>(N=62) | Pandemic<br>Cohort<br>(n=238) | Historical Cohort vs.<br>Pandemic Cohort |  |
|  |  |  |  | p† | Test<br>statistic‡ |  |  | p† | Test<br>statistic‡ |
| Maternal Characteristics |  |  |  |  |  |  |  |  |  |
| Age at Delivery (years) | 32.0<br>(18.0, 46.0) | 32.0<br>(19.0, 45.0) | 31.0<br>(19.0, 44.0) | 0.053 | 5987 | 32.0<br>(18.0, 46.0) | 32.0<br>(19.0, 45.0) | 0.25 | 6676 |
| Race |  |  |  | 0.005 | 18.33 |  |  | <0.001 | 51.04 |
| Black or African American | 43 (14.3) | 17 (13.0) | 14 (13.1) |  |  | 12 (19.4) | 31 (13.0) |  |  |
| Native American or Alaskan Native | 6 (2.0) | 1 (0.8) | 1 (0.9) |  |  | 4 (6.5) | 2 (0.8) |  |  |
| Native Hawaiian or Other Pacific Islander | 3 (1.0) | 2 (1.5) | 1 (0.9) |  |  | 0 (0) | 3 (1.3) |  |  |
| Asian or Asian American | 17 (5.7) | 13 (9.9) | 2 (1.9) |  |  | 2 (3.2) | 15 (6.3) |  |  |
| White | 104 (34.7) | 59 (45.0) | 32 (29.9) |  |  | 13 (21.0) | 91 (38.2) |  |  |
| Other or Mixed Race | 96 (32.0) | 28 (21.4) | 37 (34.6) |  |  | 31 (50.0) | 65 (27.3) |  |  |
| Declined or Unknown | 31 (10.3) | 11 (8.4) | 20 (18.7) |  |  | 0 (0) | 31 (13.0) |  |  |
| Ethnicity |  |  |  | <0.001 | 21.07 |  |  | <0.001 | 37.85 |
| Hispanic or Latino/a/x or Spanish | 168 (56.0) | 51 (38.9) | 69 (64.5) |  |  | 48 (77.4) | 120 (50.4) |  |  |
| Not Hispanic or Latino/a/x or Spanish | 115 (38.3) | 73 (55.7) | 28 (26.2) |  |  | 14 (22.6) | 101 (42.4) |  |  |
| Declined or Unknown | 17 (5.7) | 7 (5.3) | 10 (9.3) |  |  | 0 (0) | 17 (7.1) |  |  |
| Education Attainment |  |  |  | 0.001 | 21.80 |  |  | <0.001 | 35.17 |
| Some High School | 28 (9.3) | 8 (6.1) | 9 (8.4) |  |  | 11 (17.7) | 17 (7.1) |  |  |
| High School Degree/GED/Trade School | 43 (14.3) | 10 (7.6) | 20 (18.7) |  |  | 13 (21.0) | 30 (12.6) |  |  |
| Some College | 30 (10.0) | 11 (8.4) | 11 (10.3) |  |  | 8 (12.9) | 22 (9.2) |  |  |
| 2-Year College Degree | 33 (11.0) | 10 (7.6) | 18 (16.8) |  |  | 5 (8.1) | 28 (11.8) |  |  |
| 4-Year College Degree | 79 (26.3) | 37 (28.2) | 30 (28.0) |  |  | 12 (19.4) | 67 (28.2) |  |  |
| Graduate Degree | 83 (27.7) | 53 (40.5) | 18 (16.8) |  |  | 12 (19.4) | 71 (29.8) |  |  |
| Declined or Unknown | 4 (1.3) | 2 (1.5) | 1 (0.9) |  |  | 1 (1.6) | 3 (1.3) |  |  |
| Infant Characteristics |  |  |  |  |  |  |  |  |  |
| Vaginal Delivery | 186 (62.0) | 81 (61.8) | 79 (73.8) | 0.068 | 3.32 | 26 (41.9) | 160 (67.2) | <0.001 | 16.95 |
| Primiparous | 142 (47.3) | 70 (53.4) | 49 (45.8) | 0.30 | 1.09 | 23 (37.1) | 119 (50.0) | 0.30 | 1.09 |
| Gestational Age at Birth (weeks) | 39.1 (37.0, 41.6) | 39.1 (37.0, 41.6) | 39.1 (37.0, 41.1) | 0.46 | 7396 | 39.1 (37.0, 41.0) | 39.1 (37.0, 41.6) | 0.17 | 8211 |
| Female Sex | 129 (43.0) | 49 (37.4) | 54 (50.5) | 0.059 | 3.58 | 26 (41.9) | 103 (43.3) | 0.13 | 4.14 |

<sup>†</sup>Comparisons performed using Chi Squared Test for binomial variables and Wilcoxon Rank-Sum test for continuous variables. All statistical tests were two-sided.

### Maternal SARS-CoV-2 infection during pregnancy is not associated with differences in ASQ-3 scores at 6-months

There were no significant group differences between exposed and unexposed infants in any of the five ASQ-3 subdomain scores (communication, gross motor, fine motor, problem solving, or personal-social) at 6-months (Figure 1A) in either minimally or fully adjusted models (Table 2). The majority of mothers experienced asymptomatic (36%) or mild disease (61%) and were infected in the second (48%) or third (33%) trimester (Supplemental Table S1). A small proportion of mothers had severe disease (4%) and 20% were infected in the first trimester. Neither symptom severity nor timing of infection showed an association to any ASQ-3 subdomain score in either minimally or fully adjusted models (Supplemental Table S2 and S3).

**Figure 1.**
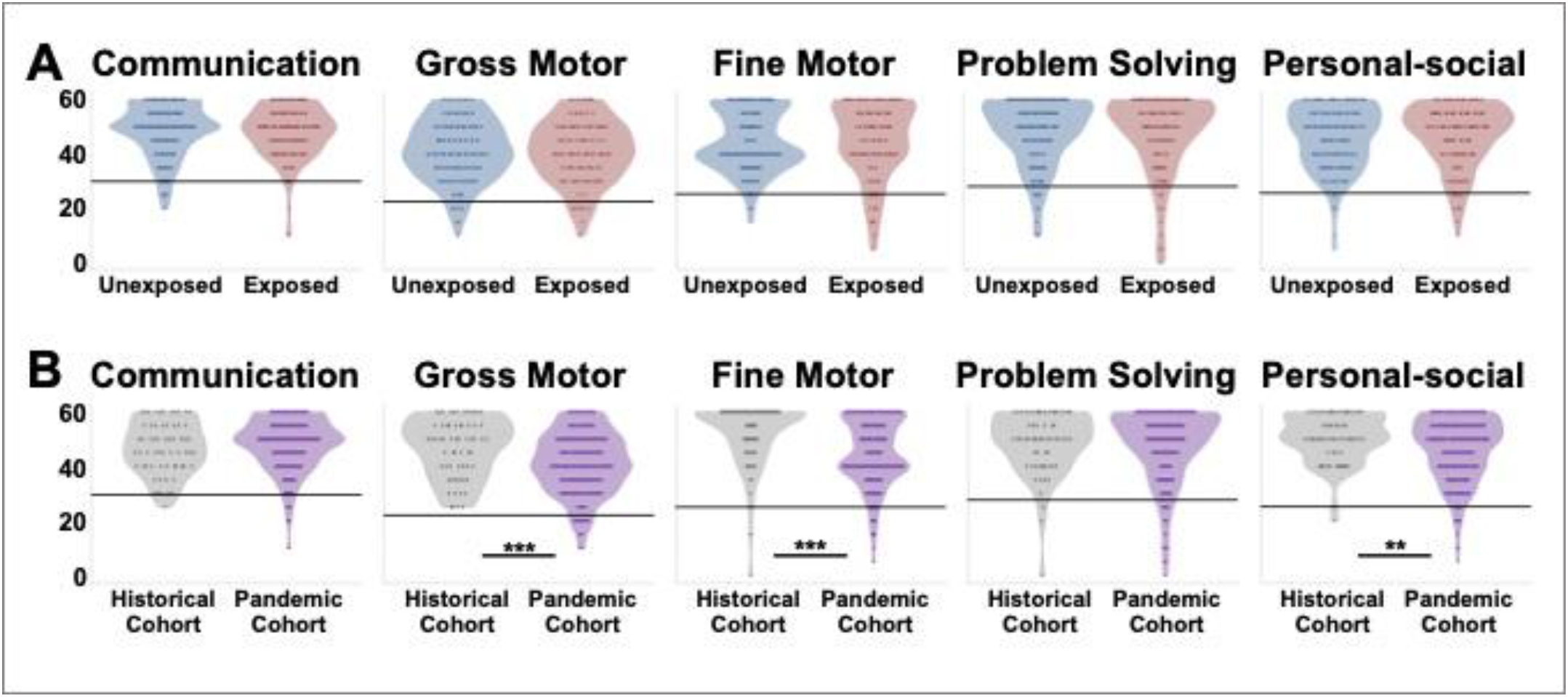
ASQ-3 scores at 6-months of age. **(A)** Total raw scores on each ASQ-3 domain for infants with *in utero* exposure to maternal SARS-CoV-2 infection (red) versus unexposed infants (blue). No significant group differences were identified on any of the ASQ-3 domains in either minimally or fully adjusted models. **(B)** Total raw scores on each ASQ-3 domain for infants born during the pandemic (purple) versus a historical cohort (gray) born at the same hospital. Gray horizontal lines represent the cutoff indicating possible delay on each subscale (communication = 29.65; gross motor = 22.25; fine motor = 25.14; problem solving = 27.72; personal-social = 25.34). *p<0.05, **p<0.01, ***p<0.001, on fully adjusted models.

**Table 2.** Comparison of 6-month ASQ-3 scores across study groups.

| ASQ-3 Subdomain | Overall Sample (N=300) | Unexposed (N=131) | Exposed (N=107) | SARS-CoV-2 Unexposed vs. Exposed |  |  |  | Historical Cohort (N=62) | Pandemic Cohort (N=238) | Historical Cohort vs. Pandemic Cohort |  |  |  |
| --- | --- | --- | --- | --- | --- | --- | --- | --- | --- | --- | --- | --- | --- |
|  |  |  |  | Minimally Adjusted |  | Fully Adjusted |  |  |  | Minimally Adjusted |  | Fully Adjusted |  |
|  |  |  |  | F (DF) | p | F (DF) | p |  |  | F (DF) | p | F (DF) | p |
| Mean Score (SD) |  |  |  |  |  |  |  |  |  |  |  |  |  |
| Communication | 47.7 (9.70) | 48.1 (9.76) | 47.7 (9.77) | 0.07 (1,233) | 0.78 | 0.07 (1,216) | 0.78 | 46.8 (9.54) | 47.9 (9.74) | 0.66 (1,295) | 0.41 | 0.68 (1,278) | 0.41 |
| Gross Motor | 42.3 (11.6) | 41.5 (11.4) | 40.7 (11.8) | 0.29 (1,233) | 0.58 | 0.29 (1,216) | 0.58 | 46.8 (10.8) | 41.1 (11.6) | 12.83 (1,295) | <0.001 | 12.63 (1,278) | <0.001 |
| Fine Motor | 46.5 (12.7) | 45.0 (11.1) | 45.1 (14.0) | 0.004 (1,233) | 0.94 | 0.004 (1,216) | 0.94 | 51.9 (12.1) | 45.1 (12.5) | 15.00 (1,295) | <0.001 | 15.39 (1,278) | <0.001 |
| Problem Solving | 47.8 (13.0) | 47.8 (12.3) | 47.3 (14.5) | 0.07 (1,233) | 0.78 | 0.07 (1,216) | 0.78 | 48.6 (11.6) | 47.6 (13.3) | 0.33 (1,295) | 0.56 | 0.33 (1,278) | 0.56 |
| Personal-Social | 47.2 (11.2) | 46.0 (11.5) | 46.7 (11.6) | 0.23 (1,233) | 0.62 | 0.24 (1,216) | 0.62 | 50.6 (8.78) | 46.3 (11.6) | 7.60 (1,295) | 0.006 | 7.60 (1,278) | 0.006 |
| Below Cutoff (%) |  |  |  |  |  |  |  |  |  | Fisher's Exact Test<sup>†</sup> p |  |  |  |
| Communication | 10 (3.3) | 6 (4.6) | 3 (2.8) |  |  |  |  | 1 (1.6) | 9 (3.8) | 0.35 |  |  |  |
| Gross Motor | 18 (6.0) | 8 (6.1) | 10 (9.3) |  |  |  |  | 0 (0) | 18 (7.6) | 0.01 |  |  |  |
| Fine Motor | 22 (7.3) | 5 (3.8) | 14 (13.1) |  |  |  |  | 3 (4.8) | 19 (8.0) | 0.30 |  |  |  |
| Problem Solving | 23 (7.7) | 10 (7.6) | 10 (9.3) |  |  |  |  | 3 (4.8) | 20 (8.4) | 0.26 |  |  |  |
| Personal-Social | 16 (5.3) | 6 (4.6) | 8 (7.5) |  |  |  |  | 2 (3.2) | 14 (5.9) | 0.32 |  |  |  |
| Any domain | 53 (17.7) | 22 (16.8) | 24 (22.4) |  |  |  |  | 7 (11.3) | 46 (19.3) | 0.10 |  |  |  |

<sup>†</sup>Fisher's Exact Tests were one-sided to test whether a greater proportion of infants in the pandemic cohort met the ASQ-3 cutoffs compared to infants in the historical cohort.

### Birth during the pandemic is associated with lower ASQ-3 scores in several domains at 6-months

Our primary analysis revealed no associations between SARS-CoV-2 status, timing, or severity and ASQ-3 scores. Therefore, exposed and unexposed infants in the pandemic cohort were pooled together and compared to infants in the historical cohort. Compared to those in the historical cohort, infants in the pandemic cohort had significantly lower mean scores on the gross motor (mean difference -5.68, 95% CI [-8.80, -2.56], F1, 295=12.83, *p*=0.0003, n^2^=0.016), fine motor (mean difference -6.77, 95% CI [-10.21, -3.33], F1, 295=15.00, *p*=0.0001, n^2^=0.017), and personal-social (mean difference -4.21, 95% CI [-7.23, -1.20], F1, 295=7.60, *p*=0.006, n^2^=0.008) subdomains in the minimally adjusted models (Figure 1B, Table 2, Supplemental Table S4). These group differences persisted in the fully adjusted model (gross motor: (mean difference -5.68, 95% CI [-8.82, -2.53], F1, 278=12.63, *p*= 0.0004, n^2^= 0.008), fine motor: (mean difference -6.77, 95% CI [-10.16, -3.37], F1, 278=15.39, *p*=0.0001, n^2^= 0.011), personal-social: (mean difference -4.21, 95% CI [-7.23, -1.20], F1, 278=7.60, *p*=0.006, n^2^= 0.005)). Sensitivity analyses excluding five infants diagnosed with COVID-19 between birth and ASQ-3 assessments did not alter findings (Supplemental Table S5 and S6). Sensitivity analyses excluding 15 infants in the historical cohort who were assessed during the pandemic did not alter findings on fine and gross motor scores, but personal-social scores no longer differed significantly between cohorts (Supplemental Table S7).

Though not specifically powered to detect differences in the proportion of infants who met screening cutoffs for delay, a greater proportion of infants in the pandemic cohort met the gross motor cutoff (one-sided Fisher’s, *p*=0.01) (Table 2).

### First trimester of pregnancy during the height of the pandemic is associated with lowest ASQ-3 personal-social scores at 6-months

We investigated whether the timing of pregnancy relative to the peak of the pandemic in NYC was associated with scores on the ASQ-3 subdomains that were identified as different between the pandemic and historical cohorts using fully adjusted models. There was a significant effect of trimester of pregnancy on fine motor (F3, 277=6.04, *p*=0.0005, n^2^= 0.016) and gross motor scores (F3, 277=5.34, *p*=0.001, n^2^= 0.012). However, Tukey contrasts did not reach the significance threshold (*p*>.05) (Supplemental Table S8 and S9). There was also a significant effect of trimester of pregnancy on infant personal-social scores (F3, 277=4.05, *p*= 0.007, n^2^= 0.025). Compared to historic controls, infants born to mothers who were in their first trimester of pregnancy during the peak of the pandemic had a significant reduction in personal-social scores (mean difference -6.16, 95% CI [-12.22, -0.10], *p*=0.009) (Figure 2, Supplemental Table S8 and S10).

**Figure 2.**
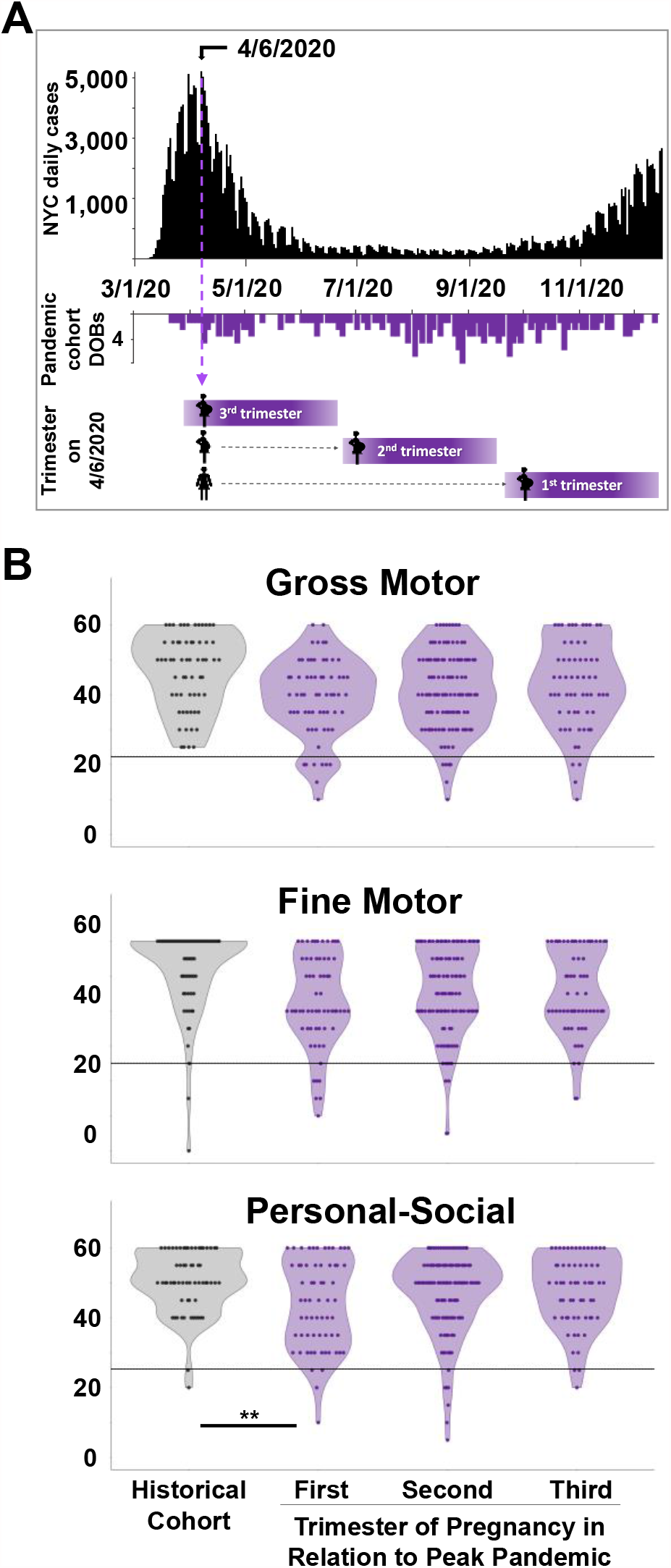
Relationship between trimester of pregnancy during the pandemic peak and ASQ-3subdomain scores at 6-months of age. **(A)** The histogram in black represents the number of recorded daily SARS-CoV-2 positive cases in NYC between March and December, 2020. The histogram in purple represents the dates of birth in two-day bins of infants enrolled in the pandemic cohort. The diagram below depicts the trimester of pregnancy corresponding to the dates of birth in the purple histogram in relation to the peak of the COVID-19 pandemic in NYC (April 6^th^, 2020). **(B)** ASQ-3 gross motor, fine motor, and personal-social scores for infants in the historical cohort versus infants in the pandemic cohort by maternal trimester of pregnancy at the peak of the pandemic in NYC (April 6^th^, 2020). Gray horizontal lines represent the cutoff indicating possible delay on each subscale (gross motor = 22.25; fine motor = 25.14; personal-social = 25.34). *p<0.05, **p<0.01, ***p<0.001, on fully adjusted model.

## Discussion

The clinical and research communities have emphasized the critical need for information regarding the neurodevelopmental effects of fetal exposure to maternal SARS-CoV-2 infection^27-31^. Prior studies of the association between fetal exposure to maternal viral infections, MIA, and atypical child neurodevelopment, led to the speculation that maternal SARS-CoV-2 infection during pregnancy may be associated with global developmental delays or specific neurodevelopmental disorders^27-31^. To our knowledge, our analysis is the first to examine this association and, contrary to the proposed hypothesis, we did not find an association between maternal SARS-CoV-2 infection status, timing, or severity and infant neurodevelopment at 6-months of age as measured using a standardized screener. However, infants born during the pandemic, regardless of maternal SARS-CoV-2 status, scored significantly lower on the gross motor, fine motor, and personal-social subdomains of the ASQ-3 when compared to a historical cohort of infants born at the same institution at least two months prior to the start of the pandemic. Furthermore, early gestation at the peak of the pandemic in NYC was associated with lowest personal-social scores compared to historic controls. Taken together, these findings suggest the potential for a significant public health impact on the generation born during the COVID-19 pandemic, necessitating further investigation.

Despite little evidence that coronaviruses, such as SARS-CoV, MERS-CoV, and now SARS-CoV-2, can cross the placenta^41,42^, the speculation that *in utero* SARS-CoV-2 exposure may impact infant neurobehavior was based on the established role of MIA on fetal brain development and fueled by recent findings of increased interleukin-6 (IL-6) in SARS-CoV-2-infected pregnant women^43^. In animal models of MIA, both viral exposure and direct injection of IL-6 lead to dose-dependent atypical behavior in offspring^44,45^. Although SARS-CoV-2 severity during pregnancy is positively correlated with IL-6 levels^43^, our analysis did not reveal an association between severity and ASQ-3 scores. It should be noted that the majority of our participants experienced an asymptomatic or mild SARS-CoV-2 infection, and therefore, infants exposed to significant MIA may not be represented in our data. Similarly, early gestational exposure is most likely to be associated with adverse neurodevelopmental outcomes^46-48^. Given that only 21 maternal SARS-Cov-2 infections (8 imputed) in our sample occurred in the first trimester, our results provide limited power for detecting the effects of infection at this early timepoint.

Together, the lack of neurodevelopmental differences between *in utero* SARS-CoV-2 exposed and unexposed infants and the observed group differences between the historical and pandemic cohorts suggest COVID-19-related stress should be considered as a potential underlying mechanism. Reported stressors have included job loss, food insecurity, and loss of housing,^49^ and the pandemic has resulted in significant increases in symptoms of anxiety and depression^50^. Consistent with our finding that infants born to women who were in the first trimester of pregnancy during the pandemic peak had the greatest reduction in personal-social scores, data from numerous cohort studies have demonstrated that prenatal perceived stress, loneliness, and objective stress, especially during early gestation, are associated with increased risk for adverse neurodevelopment in children^3,51-59^.

Although consistent with prior studies of children born during natural disasters, interpretation of our results should take into account several limitations. The data included in this analysis represent a single center, which may impact generalizability. Critically, it is also possible that developmental outcomes beyond 6-months of age will show different patterns as this is a relatively early timepoint. The ASQ-3 has modest agreement with objective measures of development^60,61^ and is often used by general pediatricians^62^. However, as a parent-report measure, our results may reflect parental-perception of – rather than objective differences in – infant neurodevelopment. Nonetheless, parental perception of development also has implications for long-term child outcomes^63^. Notable strengths of our study include detailed medical records on maternal SARS-CoV-2 status, timing, and severity, and availability of a historical comparison group from the same hospital center using the same neurodevelopmental assessment.

This first analysis of the neurodevelopment of infants born during the COVID-19 pandemic supports the critical need for long-term monitoring of these children to mitigate the substantial sequelae observed in generations born during previous pandemics.

## Data Availability

Data are available on request through formal submission to the COVID-19 Mother Baby Outcomes (COMBO) Study Maternal-Child Research Oversight (MACRO) Committee.

## Acknowledgements

The authors are grateful for the institutional support provided by the Maternal-Child Research Oversight (MaCRO) Committee and the Departments of Pediatrics, Psychiatry, and Obstetrics and Gynecology at Columbia University Irving Medical Center, which made possible the collection of time-sensitive information at the height of the COVID-19 pandemic prior to the availability of external funding opportunities. The authors wish to thank the entire COVID-19 Mother Baby Outcomes (COMBO) Initiative Team for their incredible collaborative and persevering contributions during uncertain times. Finally, the authors extend particular gratitude toward the families enrolled in COMBO, whose participation continues to inform our understanding of this unprecedented global event.

## SUPPLEMENTAL APPENDIX

### Power analyses

Our power analyses were based on our primary outcome, which aimed to investigate mean ASQ-3 subdomain score differences between infants with and without *in utero* exposure to maternal SARS-CoV-2 infections. The developers of the ASQ-3 recommend referral for infants and children scoring less than or equal to 2.0 standard deviations (SD) below normative means on any of the five subdomains. They further recommend monitoring and possible referral for infants and children scoring between 1.0 and 2.0 SD below normative means. Based on their published data, 6-month normative values are (mean±SD): 48.90±9.63 for communication, 45.64±11.69 for gross motor, 48.93±11.90 for fine motor, 50.41±11.30 for problem solving and 48.31±11.48 for personal-social. Using these normative values, our power analysis aimed to ensure 90% power to detect a 0.5 SD difference between groups, with alpha set to <0.05. Given COMBO’s enrollment has an approximate 1:1 ratio, achieving this requires a sample size of a minimum of 166 infants for problem solving and a minimum of 168 infants for all other subdomains. The current manuscript includes a total of 238 COMBO infants and therefore our analyses were well-powered to detect differences in mean ASQ-3 subdomain scores of at least 0.5 SD between SARS-CoV-2 exposed versus unexposed infants.

**Table S1.** Maternal SARS-CoV-2 timing and severity.

| **SARS-CoV-2 Severity**<br>No. (%) | **First Trimester** |  | **Second Trimester** |  | **Third Trimester** |  |
| --- | --- | --- | --- | --- | --- | --- |
|  | **Exact Only**<br>**(n=13)** | **Exact/**<br>**Imputed**<br>**(n=21)** | **Exact**<br>**Only**<br>**(n=34)** | **Exact/**<br>**Imputed**<br>**(n=51)** | **Exact Only**<br>**(n=32)** | **Exact/**<br>**Imputed**<br>**(n=35)** |
| **Asymptomatic** | 1 (7.7) | 9 (42.9) | 2 (5.9) | 19 (37.3) | 7 (21.9) | 10 (28.6) |
| **Mild** | 12 (92.3) | 12 (57.1) | 32 (94.1) | 32 (62.7) | 21 (65.6) | 21 (60.0) |
| **Severe** | 0 (0) | 0 (0) | 0 (0) | 0 (0) | 4 (12.5) | 4 (11.4) |

**Table S2.**
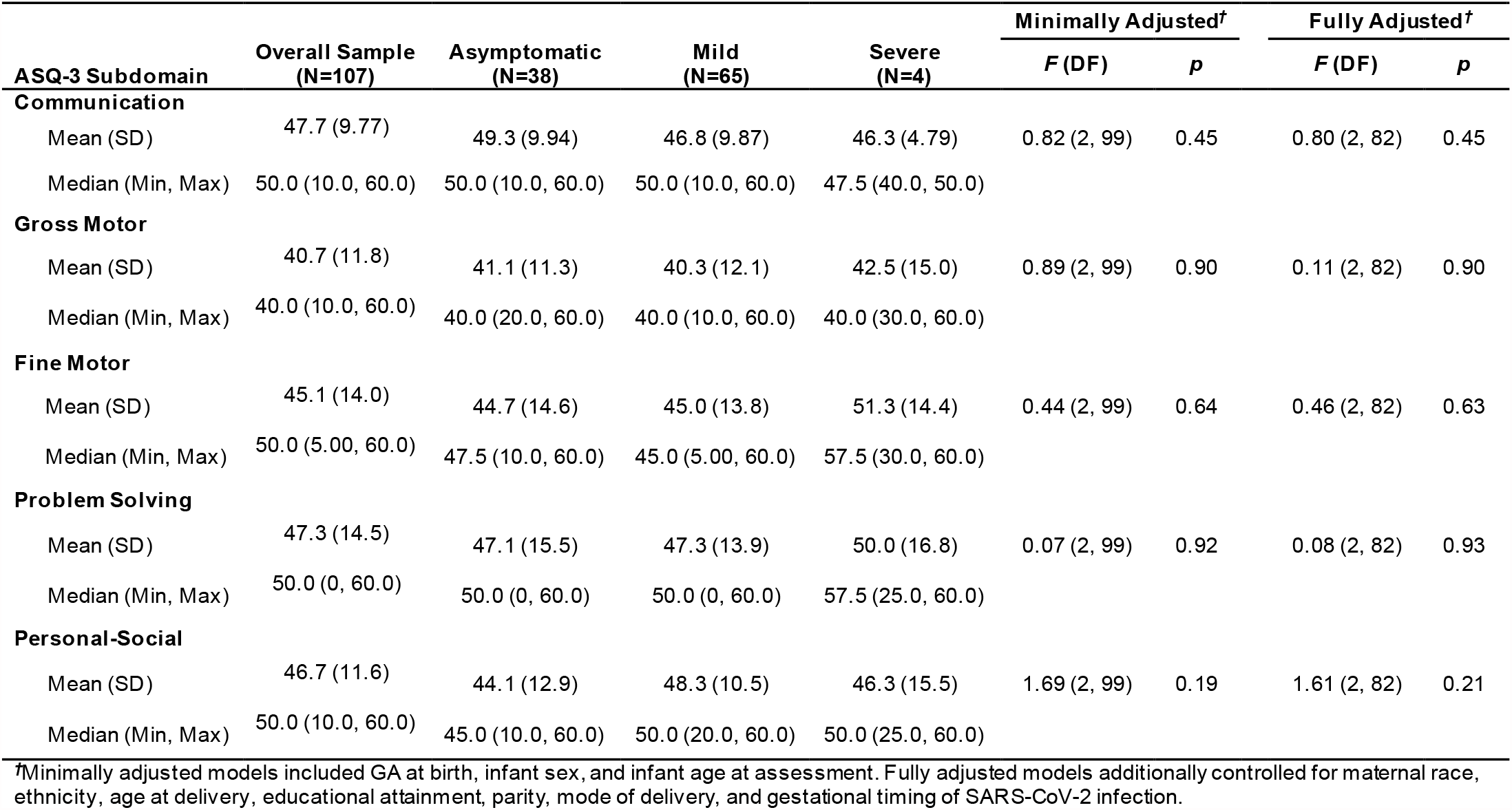
Comparison of ASQ-3 Subdomain scores across maternal SARS-CoV-2 severity.

|  |  |  |  |  |  |  |  |  |
| --- | --- | --- | --- | --- | --- | --- | --- | --- |
| ASQ-3 Subdomain | Overall Sample<br>(N=107) | Asymptomatic<br>(N=38) | Mild<br>(N=65) | Severe<br>(N=4) | Minimally Adjusted<sup>†</sup> |  | Fully Adjusted<sup>†</sup> |  |
|  |  |  |  |  | F (DF) | p | F (DF) | p |
| Communication |  |  |  |  |  |  |  |  |
| Mean (SD) | 47.7 (9.77) | 49.3 (9.94) | 46.8 (9.87) | 46.3 (4.79) | 0.82 (2, 99) | 0.45 | 0.80 (2, 82) | 0.45 |
| Median (Min, Max) | 50.0 (10.0, 60.0) | 50.0 (10.0, 60.0) | 50.0 (10.0, 60.0) | 47.5 (40.0, 50.0) |  |  |  |  |
| Gross Motor |  |  |  |  |  |  |  |  |
| Mean (SD) | 40.7 (11.8) | 41.1 (11.3) | 40.3 (12.1) | 42.5 (15.0) | 0.89 (2, 99) | 0.90 | 0.11 (2, 82) | 0.90 |
| Median (Min, Max) | 40.0 (10.0, 60.0) | 40.0 (20.0, 60.0) | 40.0 (10.0, 60.0) | 40.0 (30.0, 60.0) |  |  |  |  |
| Fine Motor |  |  |  |  |  |  |  |  |
| Mean (SD) | 45.1 (14.0) | 44.7 (14.6) | 45.0 (13.8) | 51.3 (14.4) | 0.44 (2, 99) | 0.64 | 0.46 (2, 82) | 0.63 |
| Median (Min, Max) | 50.0 (5.00, 60.0) | 47.5 (10.0, 60.0) | 45.0 (5.00, 60.0) | 57.5 (30.0, 60.0) |  |  |  |  |
| Problem Solving |  |  |  |  |  |  |  |  |
| Mean (SD) | 47.3 (14.5) | 47.1 (15.5) | 47.3 (13.9) | 50.0 (16.8) | 0.07 (2, 99) | 0.92 | 0.08 (2, 82) | 0.93 |
| Median (Min, Max) | 50.0 (0, 60.0) | 50.0 (0, 60.0) | 50.0 (0, 60.0) | 57.5 (25.0, 60.0) |  |  |  |  |
| Personal-Social |  |  |  |  |  |  |  |  |
| Mean (SD) | 46.7 (11.6) | 44.1 (12.9) | 48.3 (10.5) | 46.3 (15.5) | 1.69 (2, 99) | 0.19 | 1.61 (2, 82) | 0.21 |
| Median (Min, Max) | 50.0 (10.0, 60.0) | 45.0 (10.0, 60.0) | 50.0 (20.0, 60.0) | 50.0 (25.0, 60.0) |  |  |  |  |

<sup>†</sup>Minimally adjusted models included GA at birth, infant sex, and infant age at assessment. Fully adjusted models additionally controlled for maternal race, ethnicity, age at delivery, educational attainment, parity, mode of delivery, and gestational timing of SARS-CoV-2 infection.

**Table S3.** Comparison of ASQ-3 subdomain scores across maternal SARS-CoV-2 trimester.

| Table S3. Comparison of ASQ-3 subdomain scores across maternal SARS-CoV-2 trimester of infection |  |  |  |  |  |  |  |  |
| --- | --- | --- | --- | --- | --- | --- | --- | --- |
| ASQ-3 Subdomain | Overall Sample<br>(N=107) | First trimester<br>(N=21) | Second trimester<br>(N=51) | Third trimester<br>(N=35) | Minimally Adjusted<sup>†</sup> |  | Fully Adjusted<sup>†</sup> |  |
|  |  |  |  |  | F (DF) | p | F (DF) | p |
| Communication |  |  |  |  |  |  |  |  |
| Mean (SD) | 47.7 (9.77) | 45.5 (11.2) | 48.8 (9.83) | 47.4 (8.78) | 0.94 (2, 99) | 0.39 | 0.92 (2, 82) | 0.40 |
| Median (Min, Max) | 50.0 (10.0, 60.0) | 45.0 (10.0, 60.0) | 50.0 (10.0, 60.0) | 50.0 (20.0, 60.0) |  |  |  |  |
| Gross Motor |  |  |  |  |  |  |  |  |
| Mean (SD) | 40.7 (11.8) | 36.2 (13.1) | 41.8 (10.5) | 41.7 (12.5) | 2.07 (2, 99) | 0.13 | 2.08 (2, 82) | 0.13 |
| Median (Min, Max) | 40.0 (10.0, 60.0) | 35.0 (10.0, 55.0) | 40.0 (10.0, 60.0) | 40.0 (15.0, 60.0) |  |  |  |  |
| Fine Motor |  |  |  |  |  |  |  |  |
| Mean (SD) | 45.1 (14.0) | 41.2 (17.2) | 46.1 (13.9) | 46.1 (11.9) | 1.04 (2, 99) | 0.35 | 1.10 (2, 82) | 0.34 |
| Median (Min, Max) | 50.0 (5.00, 60.0) | 40.0 (5.00, 60.0) | 50.0 (5.00, 60.0) | 50.0 (15.0, 60.0) |  |  |  |  |
| Problem Solving |  |  |  |  |  |  |  |  |
| Mean (SD) | 47.3 (14.5) | 45.2 (19.9) | 47.3 (13.6) | 48.7 (12.1) | 0.33 (2, 99) | 0.72 | 0.34 (2, 82) | 0.71 |
| Median (Min, Max) | 50.0 (0, 60.0) | 55.0 (0, 60.0) | 50.0 (0, 60.0) | 50.0 (15.0, 60.0) |  |  |  |  |
| Personal-Social |  |  |  |  |  |  |  |  |
| Mean (SD) | 46.7 (11.6) | 44.0 (15.3) | 47.6 (10.4) | 47.0 (10.9) | 0.66 (2, 99) | 0.52 | 0.64 (2, 82) | 0.53 |
| Median (Min, Max) | 50.0 (10.0, 60.0) | 50.0 (10.0, 60.0) | 50.0 (15.0, 60.0) | 50.0 (20.0, 60.0) |  |  |  |  |
| †Minimally adjusted models included GA at birth, infant sex, and infant age at assessment. Fully adjusted models additionally controlled for maternal race, ethnicity, age at delivery, educational attainment, parity, mode of delivery, and gestational timing of SARS-CoV-2 infection. |  |  |  |  |  |  |  |  |

**Table S4.** Estimated marginal means of significant effects.

| **ASQ-3 Subdomain**<br>Estimated Marginal Mean ± SE (95% CI) | **Minimally Adjusted** | | **Fully Adjusted** | |
| --- | --- | --- | --- | --- |
|  | **Historical Cohort**<br>**(N=62)** | **Pandemic Cohort**<br>**(n=238)** | **Historical Cohort**<br>**(N=62)** | **Pandemic Cohort**<br>**(n=238)** |
| **Gross Motor** | 45.1 ± 1.53<br>(42.1, 48.1) | 41.3 ± 0.74<br>(39.8, 42.7) | 45.5 ± 2.32<br>(40.9, 50.1) | 41.9 ± 1.82<br>(38.4, 45.5) |
| **Fine Motor** | 49.9 ± 1.68<br>(46.6, 53.2) | 45.5 ± 0.81<br>(43.9, 47.1) | 50.0 ± 2.51<br>(45.0, 54.9) | 45.5 ± 1.97<br>(41.6, 49.4) |
| **Personal-Social** | 49.6 ± 1.47<br>(46.7, 52.5) | 46.8 ± 0.716<br>(45.4, 48.3) | 47.8 ± 2.22<br>(43.4, 52.2) | 45.0 ± 1.75<br>(41.6, 48.4) |

**Table S5.** Infant postnatal SARS-CoV-2 history.

| **Variable**<br>No. (%) | **Overall**<br>**Pandemic**<br>**Sample**<br>**(N=237)** | **SARS-CoV-2**<br>**Unexposed**<br>**(N=130)** | **SARS-CoV-2**<br>**Exposed**<br>**(N=107)** |
| --- | --- | --- | --- |
| **Result of Infant SARS-CoV-2 Test** |  |  |  |
| Positive | 4 (1.7) | 3 (2.3) | 1 (0.9) |
| Negative | 71 (30.0) | 16 (12.3) | 55 (51.4) |
| Other | 3 (1.3) | 2 (1.5) | 1 (0.9) |
| Not tested | 159 (67.1) | 109 (83.8) | 50 (46.7) |
| **Infant Diagnosed with SARS-CoV-2** |  |  |  |
| Yes | 5 (2.1) | 4 (3.1) | 1 (0.9) |
| No | 231 (97.5) | 125 (96.2) | 106 (99.1) |
| Prefer not to answer | 1 (0.4) | 1 (0.8) | 0 (0) |

**Table S6.**
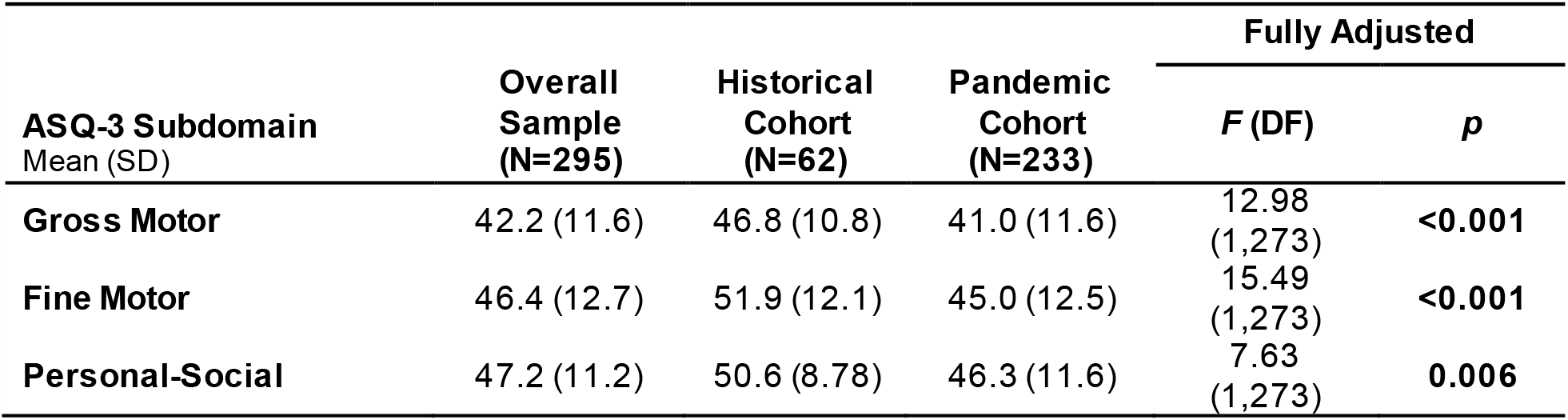
Comparison of ASQ-3 subdomain scores between historical and pandemic cohorts after excluding 5 infants from the pandemic cohort who tested positive for SARS-CoV-2 or were diagnosed with COVID-19.

| ASQ-3 Subdomain<br>Mean (SD) | Overall<br>Sample<br>(N=295) | Historical<br>Cohort<br>(N=62) | Pandemic<br>Cohort<br>(N=233) | Fully Adjusted |  |
| --- | --- | --- | --- | --- | --- |
|  |  |  |  | *F* (DF) | *p* |
| **Gross Motor** | 42.2 (11.6) | 46.8 (10.8) | 41.0 (11.6) | 12.98<br>(1,273) | **<0.001** |
| **Fine Motor** | 46.4 (12.7) | 51.9 (12.1) | 45.0 (12.5) | 15.49<br>(1,273) | **<0.001** |
| **Personal-Social** | 47.2 (11.2) | 50.6 (8.78) | 46.3 (11.6) | 7.63<br>(1,273) | **0.006** |

**Table S7.** Comparison of ASQ-3 subdomain scores between historical and pandemic cohorts after excluding 15 infants from the historical cohort assessed during the pandemic.

| ASQ-3 Subdomain<br>Mean (SD) | Overall<br>Sample<br>(N=300) | Historical<br>Cohort<br>(N=47) | Pandemic<br>Cohort<br>(N=238) | Fully Adjusted |  |
| --- | --- | --- | --- | --- | --- |
|  |  |  |  | *F* (DF) | *p* |
| **Gross Motor** | 41.9 (11.6) | 45.7 (11.4) | 41.1 (11.6) | 6.76 | **0.009** |
| **Fine Motor** | 46.2 (12.6) | 52.1 (11.9) | 45.1 (12.5) | 13.48 | **<0.001** |
| **Personal-Social** | 46.8 (11.2) | 48.9 (9.03) | 46.3 (11.6) | 2.23 | 0.13 |

**Table S8.** Tukey Pairwise Contrasts: trimester of pregnancy relative to peak SARS-CoV-2 cases in NYC. (fully adjusted models only)

| Domain and Contrast | Difference Estimate ± Standard Error | Tukey t-statistic | Tukey p-value | 95% CI of difference |
| --- | --- | --- | --- | --- |
| **Gross Motor** |  |  |  |  |
| Trimester 1 – Historical Controls | -4.95 ± 2.67 | -1.86 | 0.06 | 11.82, 1.91 |
| Trimester 2 – Historical Controls | -3.00 ± 2.45 | -1.23 | 0.22 | -9.29, 3.28 |
| Trimester 3 – Historical Controls | -3.26 ± 2.64 | -1.23 | -0.23 | -10.05, 3.534 |
| Trimester 2 – Trimester 1 | 1.96 ± 1.86 | 1.05 | 0.30 | -2.83, 6.74 |
| Trimester 3 – Trimester 1 | 1.70 ± 2.23 | 0.76 | 0.45 | -4.04, 7.44 |
| Trimester 3 – Trimester 2 | -0.25 ± 1.91 | -0.13 | 0.90 | -5.17, 4.67 |
| **Fine Motor** |  |  |  |  |
| Trimester 1 – Historical Controls | -5.57 ± 2.88 | -1.93 | 0.05 | -12.99, 1.85 |
| Trimester 2 – Historical Controls | -3.45 ± 2.64 | -1.31 | 0.19 | -10.24, 3.34 |
| Trimester 3 – Historical Controls | -5.19 ± 2.85 | -1.82 | 0.07 | -12.53, 2.15 |
| Trimester 2 – Trimester 1 | 2.12 ± 2.01 | 1.06 | 0.29 | -3.05, 7.28 |
| Trimester 3 – Trimester 1 | 0.38 ± 2.41 | 0.16 | 0.88 | -5.83, 6.58 |
| Trimester 3 – Trimester 2 | -1.74 ± 2.06 | -0.84 | 0.40 | -7.05, 3.57 |
| **Personal-Social** |  |  |  |  |
| Trimester 1 – Historical Controls | -6.16 ± 2.35 | -2.62 | **0.009** | -12.22, -0.10 |
| Trimester 2 – Historical Controls | -4.20 ± 2.21 | -1.90 | 0.06 | -9.90, 1.50 |
| Trimester 3 – Historical Controls | -3.46 ± 2.53 | -1.37 | 0.17 | -9.96, 3.04 |
| Trimester 2 – Trimester 1 | 1.96 ± 1.78 | 1.10 | 0.27 | -2.61, 6.54 |
| Trimester 3 – Trimester 1 | 2.70 ± 2.04 | 1.33 | 0.19 | -2.56, 7.96 |
| Trimester 3 – Trimester 2 | 0.74 ± 1.79 | 0.41 | 0.68 | -3.86, 5.34 |

**Table S9.** Raw ASQ-3 Subdomain scores by trimester of pregnancy relative to the pandemic.

| ASQ-3 Subdomain | Historic Controls<br>(N=62) | Trimester of pregnancy relative to peak SARS-CoV-2 cases in New York City (4/6/2020) |  |  |
| --- | --- | --- | --- | --- |
|  |  | Trimester 1<br>(N=64) | Trimester 2<br>(N=112) | Trimester 3<br>(N=62) |
| Communication |  |  |  |  |
| Mean (SD) | 46.8 (9.54) | 47.0 (9.99) | 48.4 (10.0) | 47.9 (8.99) |
| Median [Min, Max] | 45.0 [25.0, 60.0] | 50.0 [10.0, 60.0] | 50.0 [10.0, 60.0] | 50.0 [20.0, 60.0] |
| Gross Motor |  |  |  |  |
| Mean (SD) | 46.8 (10.8) | 39.1 (11.2) | 41.3 (11.2) | 42.8 (12.4) |
| Median [Min, Max] | 50.0 [25.0, 60.0] | 40.0 [10.0, 60.0] | 40.0 [10.0, 60.0] | 42.5 [10.0, 60.0] |
| Fine Motor |  |  |  |  |
| Mean (SD) | 51.9 (12.1) | 43.0 (12.9) | 45.8 (12.3) | 46.0 (12.1) |
| Median [Min, Max] | 60.0 [0, 60.0] | 40.0 [10.0, 60.0] | 45.0 [5.00, 60.0] | 45.0 [15.0, 60.0] |
| Problem Solving |  |  |  |  |
| Mean (SD) | 48.6 (11.6) | 45.2 (14.7) | 47.8 (13.4) | 49.7 (11.3) |
| Median [Min, Max] | 50.0 [0, 60.0] | 50.0 [0, 60.0] | 50.0 [0, 60.0] | 55.0 [15.0, 60.0] |
| Personal-Social |  |  |  |  |
| Mean (SD) | 50.6 (8.78) | 43.9 (12.5) | 46.9 (11.6) | 47.8 (10.2) |
| Median [Min, Max] | 50.0 [20.0, 60.0] | 45.0 [10.0, 60.0] | 50.0 [5.00, 60.0] | 50.0 [20.0, 60.0] |

**Table S10.**
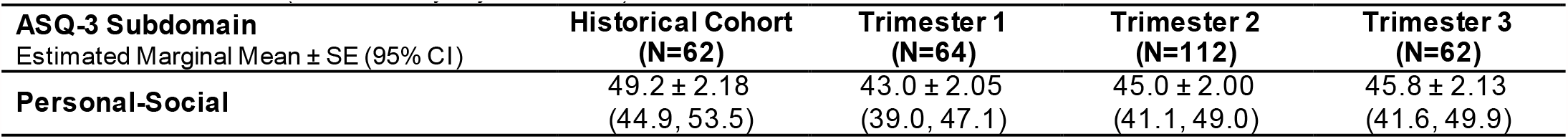
Estimated marginal means of personal-social scores by trimester of pregnancy relative to peak SARS-CoV-2 cases in NYC. (based on fully adjusted model)

